# Using Machine Learning Algorithms to predict sepsis and its stages in ICU patients

**DOI:** 10.1101/2022.03.15.22271655

**Authors:** Nimrah Ghias, Shan Ul Haq, Huzifa Arshad, Haseeb Sultan, Farhan Bashir, Syed Ameer Ghaznavi, Maria Shabbir, Yasmin Badshah, Mehak Rafiq

## Abstract

Sepsis is blood poisoning disease that occurs when body shows dysregulated host response to an infection and cause organ failure or tissue damage which may increase the mortality rate in ICU patients. As it becomes major health problem, the hospital cost for treatment of sepsis is increasing every year. Different methods have been developed to monitor sepsis electronically, but it is necessary to predict sepsis as soon as possible before clinical reports or traditional methods, because delayed in treatment can increase the risk of mortality with every single hour. For the early detection of sepsis, specifically in ICU patients, different machine learning models i.e Linear learner, Multilayer perceptron neural networks, Random Forest, lightgbm and Xgboost has trained on the data set proposed by Physio Net/ Computing in Cardiology Challenge in 2019. This study shows that Machine learning algorithms can accurately predict sepsis at the admission time of patient in ICU by using six vitals signs extracted from patient records over the age of 18 years. After comparative analysis of machine learning models, Xgboost model achieved a highest accuracy of 0.98, precision of 0.97, and recall 0.98 under the precision-recall curve on the publicly available data. Early prediction of sepsis can help clinicians to implement supportive treatments and reduce the mortality rate as well as healthcare expenses.

## INTRODUCTION

Sepsis is clinical syndrome caused by body’s overwhelming which leads to tissue damage and organ failure (9). In recent decades, sepsis still one of the life threatening disease in hospitals. It is most commonly manifested by systemic bacterial infection that involves in the production of endotoxin, but it can also be caused by fungal or viral etiology (4).It is linked with high morbidity rate, mortality rate and responsible for hospital cost (12). Globally, an estimated 30 million people diagnosed sepsis in Intensive Care Units and 6 million people died from sepsis each year (19). It is highly affected on adults and children. The pathophysiological pathways of sepsis is very complex therefore it has variety of signs and symptoms which are not easily detectable (15). The latest studies proposed that the mortality is increased with every hour if the antibiotic treatment is delayed, because some patients having sepsis even at the time of admission. Identifying risk factors earlier and commencing appropriate monitoring, before to any clinical symptoms, would have a major influence on overall mortality and financial burden of sepsis (10). Currently, available screening methods for sepsis i.e. systemic inflammatory response syndrome (SIRS), modified early warning systems (MEWS), qSOFA etc are not enough for clear identification of sepsis (8). Many researchers are concentrated on machine learning approaches for the excellent outcome and high accuracy which is superior to the every disease severity scoring systems. Basically, machine learning aims to develop algorithm that can learn and create models for prediction and data analysis which give rapid outcomes (3). This current work was designed to adopt a real time machine learning algorithms linear learner, xgboost, multilayer perceptron neural networks, lightgbm and random forest to detect sepsis at the time when patient admitted in ICU, based on PhysioNet data collected from two hospitals. In ICU, patients are admitted due to different reasons, the recognition of early sepsis with various disease states (e.g inflammation) is quite challenging because every disease in ICU shows similar instances (e.g dysregulated host response), clinical criteria (e.g change in vitals) and symptoms (e.g fever) (9). Machine learning models have ability to learn predictive patterns in data that helps to handle the complexity and wealth of digital patient data, which in turn give valid predictions about patient having sepsis. The predictive patterns can be exposed either through supervised or unsupervised learning. The algorithms that involve labeled training data (e.g. patients have sepsis or not) to predict outcomes for unforeseen data is presented as supervised learning. In contrast, the data which has no labels and determine (known and unknown) patterns in the data is included in unsupervised learning. Over the last years, many research have used a range of computational models to deal with the difficulty in prediction of sepsis at its earlier stage (9). The large number of features are retrieved from available attributes to train different machine learning models and improve their performance. After verification of the proposed algorithms, through 5-fold cross validation method build the final ensemble model is applied on public challenge database and make evaluation of this model on the hidden test set (17). The early detection of sepsis resulted in proper monitoring and management of the patient leading to significant reduction in mortality rate (15).

## METHODOLOGY

This research aims to predict sepsis at the time of patient’s admission in ICU by applying machine learning algorithms and extracted out the best model for the prediction. There are five steps involved to achieve the goal.

1. Data Description
2. Tools Used
3. Data Preprocessing
4. Feature Selection
5. Machine Learning Algorithms

### Data Description

The data is extracted from Physionet challenge 2019 which consist of 40336 PSV files, collected from two different hospitals (Training set A which involved 20336 patients of hospital A and Training set B involved 2000 patients of hospital B). Each file indicates hourly recorded data of patients after admitting in ICU. The data includes 41 variables which consists of 26 labortary values (Measure of white blood counts, Bicarbonate, etc), eight vital signs (temperature, heart rate, oxygen saturation, and systolic blood pressure etc), six demographics (gender, age, ICULOS, etc). The last variable represents sepsis label 0 and 1. 1 means the sepsis has identified in patient on the basis of sepsis 3 criteria. The data is highly imbalance that only 2932 out of 40336 patients has sepsis. Additionally, there are many variables(26 out of 41) which have missing values more than 70 percent. For early sepsis prediction, the sepsis label has shifted forward for six hours in all data (meaning that the label is set to 1 for six hours before it is officially identified).

### Tools Used

There are many machine learning libraries i.e. scikit-learn, numpy, pandas, matplotlib which are open source, and use for classification, clustering, regression and dimensionality reduction. Scikit-learn is one of the most popular libraries which is used for evaluation of model and useful to extract important features. If the dataset is highly imbalance then it is considered as quite challenging, so to deal with the imbalance dataset there is library of Imbalanced-learn which offers multiple resampling techniques i.e. SMOTE analysis.

### Data Preprocessing

It is the most important phase in data formatting and data normalization. The review of data should be carefully analyzed to avoid misleading results. Therefore, interpretation for accurate data should be done before model building. The process of data preprocessing deals with redundant.missing and noisy data and its strategies involved imputation of missing values and feature extraction. The large number of missing values in the dataset was needed to be imputed for better prediction outcomes by using different methods i.e. 0 imputation method, mean, median, mode and missforest method. The main method to normalize the data is Min Max scalar or standard scalar and then Missforest algorithm is considered as best imputation method as compared with other methods which has validated by making histogram of every variable which shows normal distribution because better imputation is the basic key for the better performance of model. Missforest can handle different kind of data i.e. continuous and categorical. This algorithm doesn’t require hyperparameter tunning. It works on the basis of Random Forest which handle all missing value according to its requirement. It predict the values on the basis of original data distribution and also useful to fix the imbalance data (20).

### Feature Selection

The mechanism of feature selection is used to filter out the most relatable features with the variable which are needed to predict. The model accuracy can be effected by using inappropriate features showing maximum outlier detection. This study has focused on six vital signs which are selected on the basis of statistical analysis by using Z test having the idea that these vital signs are present in all ICU patients and can be used for sepsis prediction. The correlation analysis has been used to extract the features that were showing highly contribution as predicting variables.

### Machine Learning Algorithms

There are many traditional methods i.e. laboratory test, qsofa score, SIRS etc. to detect sepsis but delayed in detection due to unclear symptoms cause the high mortality rate and increase the cost of hospitals therefore, there was need to predict sepsis earlier than clinical reports. For that purpose different machine learning algorithms can be used for early detection with the high sensitivity and specificity rate. For example, Xgboost, Random Forest and linear learner, LightGBM etc. Xgboost is one of the best algorithm for the classification problem and shows accurate performance. It shows iterative phenomena and combine all the results extracted from weak decision trees and gives the best prediction. In every iteration it is focused on misclassified observations. It includes the gradient boosted trees and construct the model. The prediction of sepsis was generated by using six vital signs HR, temp, O2Sat, SBP and MAP at 6hr before prediction (1). Meanwhile, XGBoost may process missing data automatically by assigning a default direction to null values. To achieve the best XGBoost model performance, evaluation of hyperparameters was required, which included number of estimators, maximum depth and learning rates. The original dataset was randomly partitioned into five subsets for this investigation. One-fold was utilized as a testing subset, while the other four-fold were used to tune the hyperparameters, with 25 percent used for calibration and the remaining 75 percent subjected to four-fold cross validation with grid search. The hyperparameters selected that have the greatest area under the receiver operator characteristic (18). Random forest was selected as the modern machine learning-based model, and it may be viewed as an extension of existing tree based classifiers which is useful for classification and regression problems. Random forest was chosen over other machine learning techniques (e.g support vector machines) because it is similar to CART and has advantages when dealing with EHR data. Random forest is an ensemble-based strategy that constructs several decision trees (i.e., “forest”) at the training data to offset the constraints of decision trees. Each tree is built from a randomly selected subset of the original training data. A random subset of the entire number of variables is evaluated at each splitting node. By adopting the mode of decision-making it can reduce the problem of overfitting (14). LightGBM is great classifier for prediction which works 6 times faster than Xgboost.It learned about those attributes which having great contribution in prediction(CHAMI et al.). It depends on histogram based algorithms which reduces consumption of memory and speed up the training step. It combines advance communication networking for parallel learning.That is why it is also known as parallel voting decision tree algorithm. In each iteration, divide the training data into multiple machines and perform a local voting decision to select the top-k attributes and a global voting decision to receive the top2k attributes(16).

Linear Learner algorithm is used for binary classification. It is having an option of normalization for preprocessing. By turning on the normalization, it moves towards the smallest sample of the data and find out mean value and standard deviation for every label and attribute. But for binary classification, only features can be normalized. There are many optimization algorithms are involved which can be used to take control for optimization processes and help to deal with hyperparameters. When many models are trained in parallel manner, then they are compared with validation set to check which model is optimal. The optimal model () gave the best F1 score () and accuracy () on the validation set. The other deep learning algorithm used for classification in advance level is multilayer perceptron neural network which is also known as feed forward neural network which involves input layer, hidden layer and output layer in which unlimited data can be used. It doesn’t only include vital signs, but also demographics or laboratory values. This algorithm doesn’t make any assumptions about distribution of data. The most attractive thing about this technique is it can trained as numerical models on new data (6). It basically consists of nodes or neurons having weights. Each neuron in MLP is connected to multiple of its neighbours, with varied weights expressing the relative importance of the various neuron inputs on the other neurons (7). The imbalanced number of neurons in hidden layer may cause the overfitting but there is no specific method to find number of neurons. It is only dependent on trail and error method (11).

## EXPERIMENTAL RESULTS

Correlation and Statistical analysis figured out those characteristics that have a significant impact on sepsis prediction.The extracted attributes with more than 70 percent of missing values has dropped and the rest of variables were used for the prediction which were imputed by using Missforest algorithm.

Training set A include number of patients 790215 and Training Set B with number of patients () and the third A/B combined dataset having patients 1552210 have used for model training by extracting six vital signs.The groups include total (684107) number of males and (868103) number of females and the age of patients which having sepsis is under 60-70 years. The summary of each dataset is presented in Fig().The data is standardized by performing SMOTE analysis give better outcomes than without SMOTE.The performance of machine learning models are summarized on these three datasets Each figure presents ROC curve of every model with the comparison of other datasets. The graphs showing area under the precision recall curve and the area operating characteristic curve of each dataset while table showing results of every model with Auc score, F1 score, precision and recall.

## DISCUSSION

Early sepsis prediction is significant problem but still challenging. This study proposed that machine learning models shows high performance on prediction (AUROC 0.98) at the spot after patient’s data entry(10). Machine learning algorithms used hourly based data after patients admitted in ICU to predict the prognosis of sepsis patients, the severity in condition of sepsis (i.e. septic shock), and maximum length of stay of septic patients in ICU. Xgboost, Random forest, Lightgbm, Linear Learner, Multilayer Perceprtron neural networks classifiers had stronger predictive power, with areas under the AUC score of 0.90, 0.92,0.94 respectively. In early stage of sepsis, usage of Random forest classifier allow to anticipate better ICU patients outcome, shows appropriate medical measures and improve the treatment which improves prognosis (17). As many biological events has happened in the pathophysiological of sepsis which leads to the disease processes and health complications. (18). Its quite difficult to deal with disease complexity in ICU and imbalance data, therefore, the advanced methods of machine learning presented the new scoring systems for accurate prediction (13). The another interesting outcome is every model trained on combined dataset Training set A and Training set B as well as on seperate datasets and showing better results on training as well as on test dataset (5).Moreover, this study also shows the importance of each feature that is having great impact on sepsis. The statistical anlaysis has been used for the purpose of validation of each attribute on the basis of Z-test. The total number of septic and non septic patients in dataset are examined and separate them in different classes and count the number of male and female having sepsis and analyze the age which is more targeted due to sepsis.The prevalence of sepsis is disproportionately higher in the elder patients and the age of a person is an independent predictor of death. The elder patients are mostly non survivors of sepsis. This analysis is good for better understanding about the data and helpful to know that sepsis mostly effects the female as compared to male. The difference in male and female shows different hormone response to an infection. The septic male and female have high estrogen level and shows the severity of illness in females than males. Females with septic shock have high anti inflammatory mediators while males have high tendency to maintain the health status. So by knowing the biological events it proved that females have severe effect towards illness than male. After the statistical and correlation analysis six vital signs has confirmed for the further process which are heart rate, temperature, oxygen saturation, respiratory rate, mean arterial pressure, systolic blood pressure and diastolic pressure. These variables having great impact in the prediction sepsis and can be used for model building. This study shows the contribution in the comparison of different machine learning models and find out the best model which can be deployed in hospitals. The model is trained on the features selected from dataset. For the prediction of sepsis, every model has presented best performance by giving ROC curve from (0.95 to 0.98). There is no limitation in distribution of features while using these models therefore, they can used to tackle the large data as well.The evaluation of predictive model occur by confusion matrix which compute the senstivity, error rate, precision and specificity while AUC is metric which differentiate the sepsis patients from other patients. In the comparison of these ensemble models, Xgboost is more preferable than random forest because Xgboost shows the integration of decision tress in sequential manner while random forest select each decision tree individually and make a random subset for construction(**?**). This model could achieve highest ROC curve because of better selection of features, dealing with imbalance data or overfitting through smote analysis was the main key for the best prediction (5).

**Figure 1.**
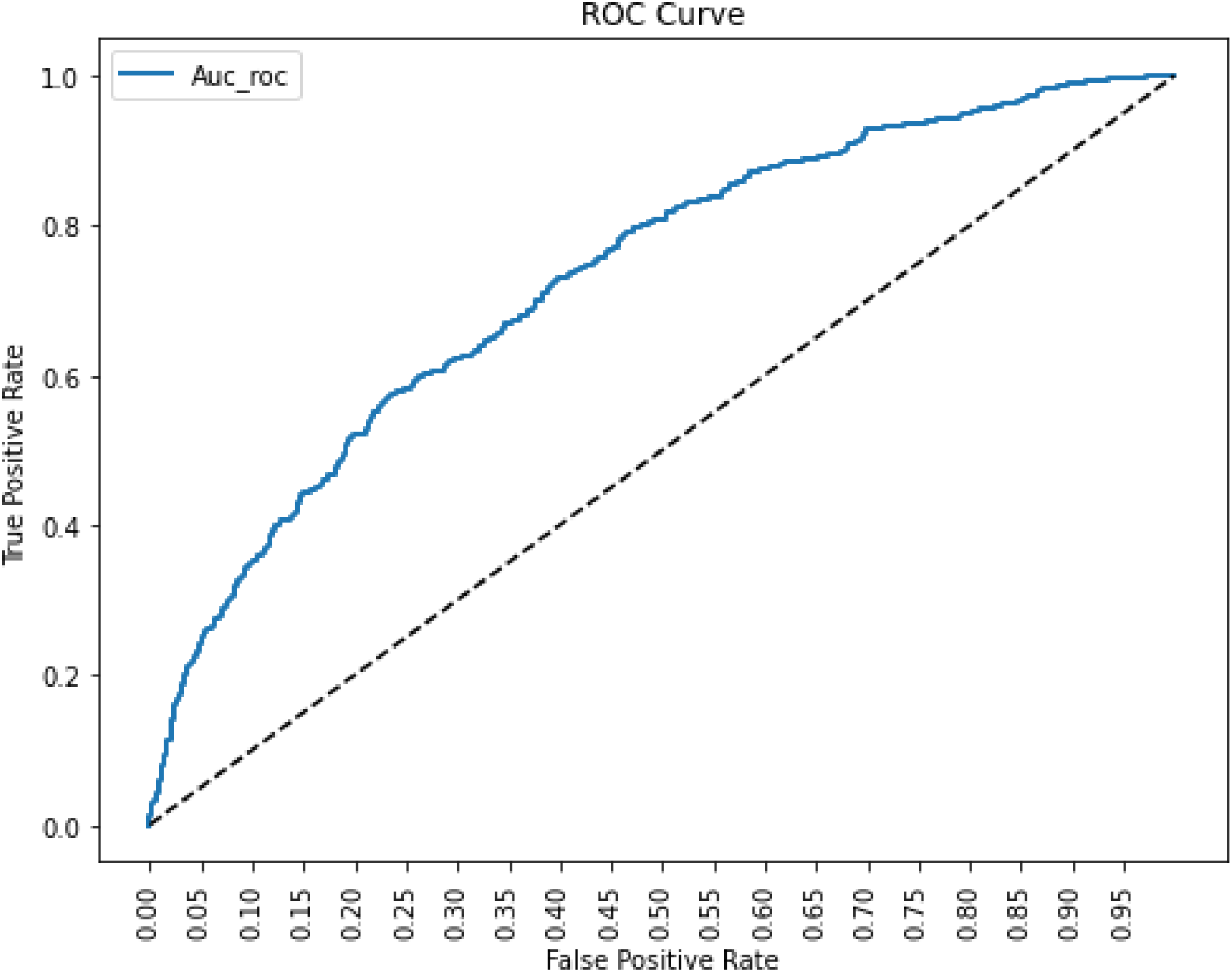
ROC curve.

**Figure 2.**
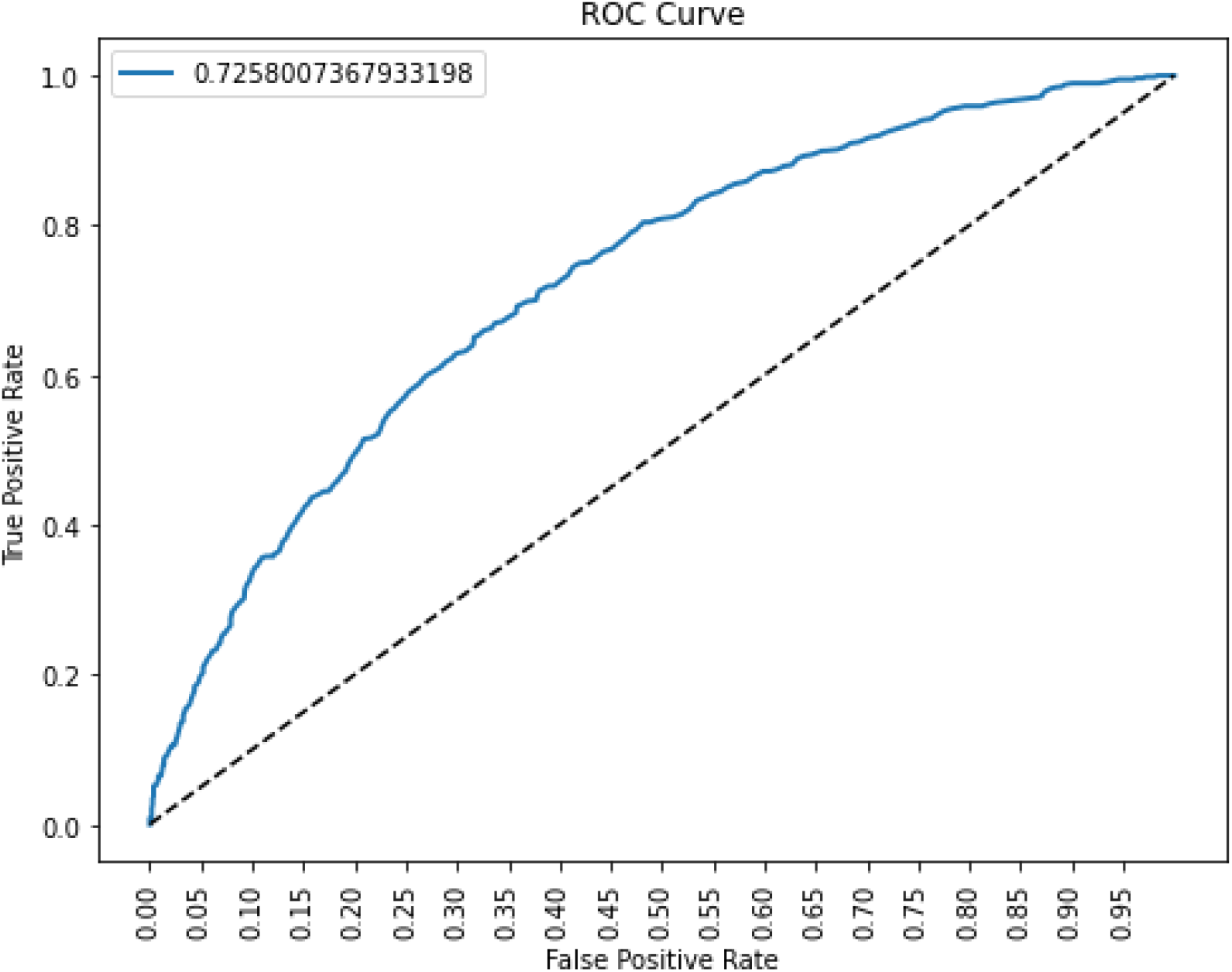
ROC curve.

**Figure 3.**
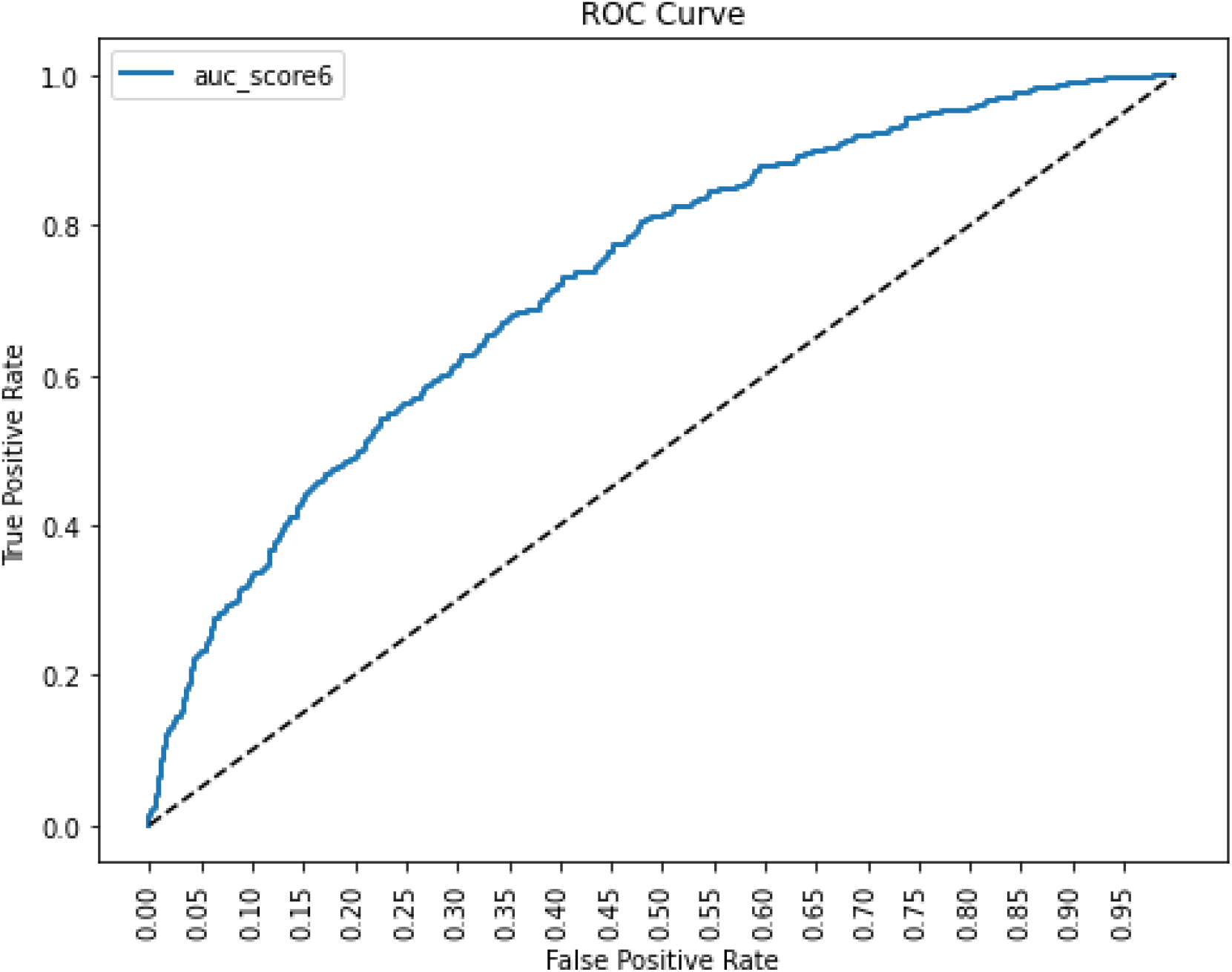
ROC curve.

**Figure 4.**
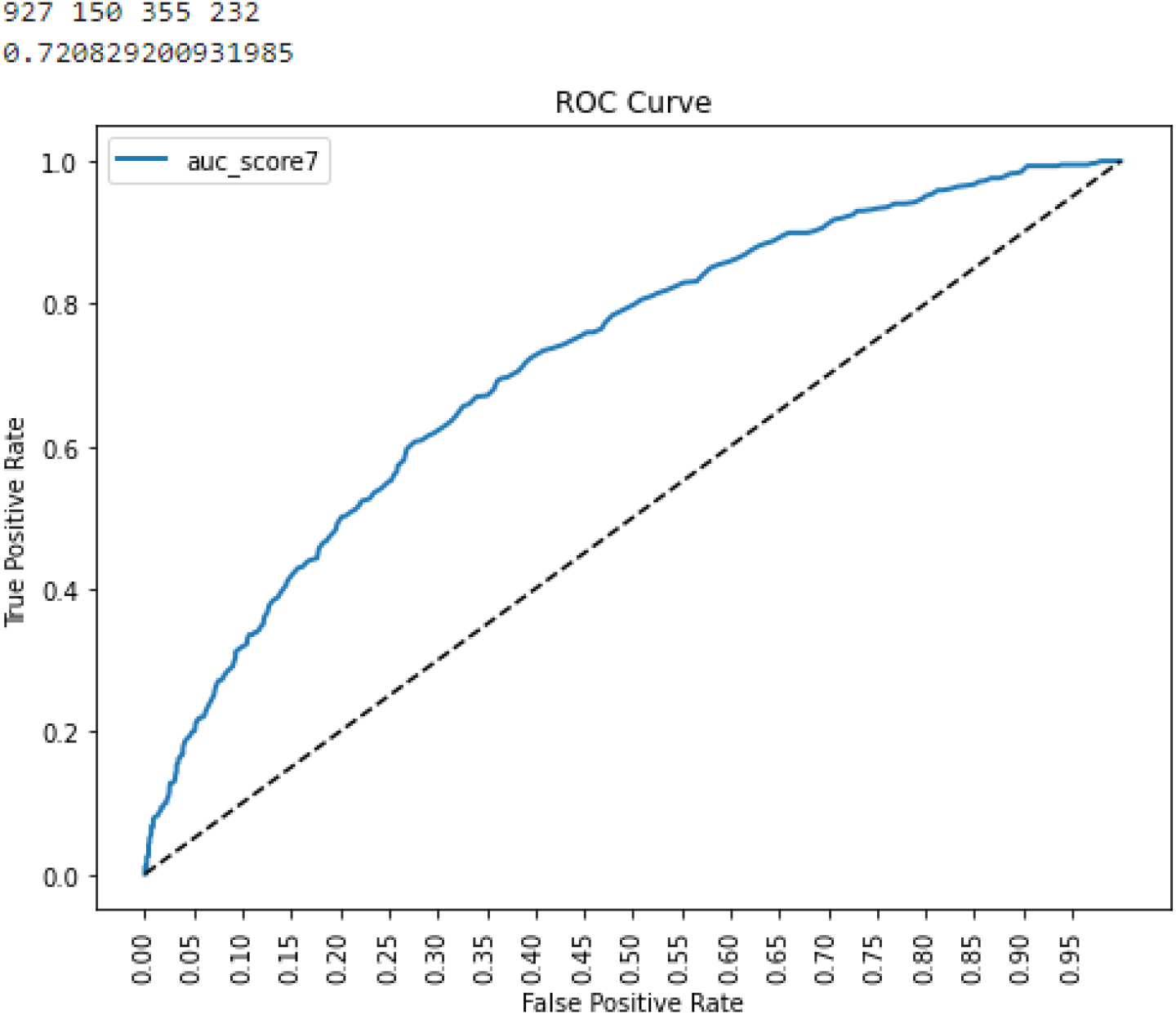
ROC curve.

## CONCLUSIONS

Sepsis is life threatening disease which cause of high mortality rate and morbidity in hospitals. Early detection is a key to overcome the death rate, therefore this study showed the development of fast and accurate machine learning algorithm for the prediction of sepsis which gives the better results than the existing scoring systems. In addition, the comparative analysis has done between five main models of machine learning by measuring their specificity and sensitivity. These models has potential to use for commercial use in ICU’s for sepsis prediction.

## Data Availability

All data produced are available online at https://physionet.org/content/challenge-2019/1.0.0/

## ACKNOWLEDGMENTS

So long and thanks for all the fish.

## REFERENCES

[1] Burdick, H., Pino, E., Gabel-Comeau, D., Gu, C., Roberts, J., Le, S., Slote, J., Saber, N., Pellegrini, E., Green-Saxena, A., et al. (2020). Validation of a machine learning algorithm for early severe sepsis prediction: a retrospective study predicting severe sepsis up to 48 h in advance using a diverse dataset from 461 us hospitals. BMC medical informatics and decision making, 20(1):1–10.

[2] Chami, S., Kaabouch, N., and Tavakolian, K. Comparative study of light-gbm and a combination of survival analysis with deep learning for early detection of sepsis.

[3] Chibani, S. and Coudert, F.-X. (2020). Machine learning approaches for the prediction of materials properties. APL Materials, 8(8):080701.

[4] Dolin, H. H., Papadimos, T. J., Chen, X., and Pan, Z. K. (2019). Characterization of pathogenic sepsis etiologies and patient profiles: a novel approach to triage and treatment. Microbiology insights, 12:1178636118825081.

[5] Fu, M., Yuan, J., Lu, M., Hong, P., and Zeng, M. (2019). An ensemble machine learning model for the early detection of sepsis from clinical data. In 2019 Computing in Cardiology (CinC), pages Page–1. IEEE.

[6] Gardner, M. W. and Dorling, S. (1998). Artificial neural networks (the multilayer perceptron)—a review of applications in the atmospheric sciences. Atmospheric environment, 32(14-15):2627–2636.

[7] Heidari, E., Sobati, M. A., and Movahedirad, S. (2016). Accurate prediction of nanofluid viscosity using a multilayer perceptron artificial neural network (mlp-ann). Chemometrics and intelligent laboratory systems, 155:73–85.

[8] Islam, M. M., Nasrin, T., Walther, B. A., Wu, C.-C., Yang, H.-C., and Li, Y.-C. (2019). Prediction of sepsis patients using machine learning approach: a meta-analysis. Computer methods and programs in biomedicine, 170:1–9.

[9] Moor, M., Rieck, B., Horn, M., Jutzeler, C. R., and Borgwardt, K. (2021). Early prediction of sepsis in the icu using machine learning: a systematic review. Frontiers in medicine, 8:348.

[10] Nemati, S., Holder, A., Razmi, F., Stanley, M. D., Clifford, G. D., and Buchman, T. G. (2018). An interpretable machine learning model for accurate prediction of sepsis in the icu. Critical care medicine, 46(4):547.

[11] Orhan, U., Hekim, M., and Ozer, M. (2011). Eeg signals classification using the k-means clustering and a multilayer perceptron neural network model. Expert Systems with Applications, 38(10):13475–13481.

[12] Parashar, A., Mohan, Y., and Rathee, N. (2021). Analysis of various health parameters for early and efficient prediction of sepsis. In IOP Conference Series: Materials Science and Engineering, volume 1022, page 012002. IOP Publishing.

[13] Su, L., Xu, Z., Chang, F., Ma, Y., Liu, S., Jiang, H., Wang, H., Li, D., Chen, H., Zhou, X., et al. (2021). Early prediction of mortality, severity, and length of stay in the intensive care unit of sepsis patients based on sepsis 3.0 by machine learning models. Frontiers in Medicine, 8:883.

[14] Taylor, R. A., Pare, J. R., Venkatesh, A. K., Mowafi, H., Melnick, E. R., Fleischman, W., and Hall, M. K. (2016). Prediction of in-hospital mortality in emergency department patients with sepsis: a local big data–driven, machine learning approach. Academic emergency medicine, 23(3):269–278.

[15] Vincent, J.-L. (2016). The clinical challenge of sepsis identification and monitoring. PloS medicine, 13(5):e1002022.

[16] Wang, D., Zhang, Y., and Zhao, Y. (2017). Lightgbm: an effective mirna classification method in breast cancer patients. In Proceedings of the 2017 International Conference on Computational Biology and Bioinformatics, pages 7–11.

[17] Yang, M., Wang, X., Gao, H., Li, Y., Liu, X., Li, J., and Liu, C. (2019). Early prediction of sepsis using multi-feature fusion based xgboost learning and bayesian optimization. In The IEEE Conference on Computing in Cardiology (CinC), volume 46, pages 1–4.

[18] Yao, R.-q., Jin, X., Wang, G.-w., Yu, Y., Wu, G.-s., Zhu, Y.-b., Li, L., Li, Y.-x., Zhao, P.-y., Zhu, S.-y., et al. (2020). A machine learning-based prediction of hospital mortality in patients with postoperative sepsis. Frontiers in Medicine, 7:445.

[19] Zabihi, M., Kiranyaz, S., and Gabbouj, M. (2019). Sepsis prediction in intensive care unit using ensemble of xgboost models. In 2019 Computing in Cardiology (CinC), pages Page–1. IEEE.

[20] Zhao, X., Shen, W., and Wang, G. (2021). Early prediction of sepsis based on machine learning algorithm. Computational Intelligence and Neuroscience, 2021.

